# Successful reboot of high-performance sporting activities by Japanese national women’s handball team in Tokyo, 2020 during the COVID-19 pandemic: An initiative by Japan Sports-Cyber Physical System (JS-CPS) of Sports Research Innovation Project (SRIP)

**DOI:** 10.1101/2021.01.29.21250745

**Authors:** Issei Ogasawara, Shigeto Hamaguchi, Ryosuke Hasegawa, Yukihiro Akeda, Naoki Ota, Gajanan S. Revankar, Shoji Konda, Takashi Taguchi, Toshiya Takanouchi, Kojiro Imoto, Nobukazu Okimoto, Katsuhiko Sakuma, Akira Uchiyama, Keita Yamasaki, Teruo Higashino, Kazunori Tomono, Ken Nakata

## Abstract

**Background:** The COVID-19 pandemic has negatively impacted sporting activities across the world. However, practical training strategies for athletes to reduce the risk of infection during the pandemic has not been definitively studied.

**Objective:** The purpose of this report was to provide an overview of our challenges encountered during the reboot of high-performance sporting activities of the Japanese national handball team during the 3rd wave of the COVID-19 pandemic in Tokyo, Japan.

**Methods:** Twenty-nine Japanese national women’s handball players and 24 staff participated in the study. To initiate the reboot of their first training camp after COVID-19 stay-home social policy, we conducted: web-based health-monitoring, SARS-CoV-2 screening with polymerase chain reaction (PCR) test, real-time automated quantitative monitoring of social distancing on-court using video-based artificial intelligence (AI) algorithm, physical intensity evaluation with wearable heart rate (HR) and acceleration sensors, and self-reported online questionnaire.

**Results:** The training camp was conducted successfully with no COVID-19 infections. The web-based health monitoring and the frequent PCR testing with short turnaround times contributed remarkably in early detection of athletes’ health problems and risk screening. During handball, the AI based on-court social-distancing monitoring revealed key time-dependent spatial metrics to define player-to-player proximity. This information facilitated positive team members’ on and off-game distancing behavior. Athletes regularly achieved around 80% of maximum HR during training, indicating anticipated improvements in achieving their physical intensities. Self-reported questionnaires related to the COVID management in the training camp revealed a sense of security among the athletes allowing them to focus singularly on their training.

**Conclusion:** The current challenge provided us considerable know-how to create and manage a safe environment for high-performing athletes in the COVID-19 pandemic via the Japan Sports-Cyber Physical System (JS-CPS) of SRIP (Japan Sports Agency, Tokyo, Japan). This report is envisioned to provide informed decisions to coaches, trainers, policymakers from the sports federations in creating targeted, infection-free, sporting and training environments.

## 1. Introduction

Physical inactivity has been recognized as one of the top four leading causes of death worldwide (1). The COVID-19 pandemic has irrevocably worsened this physical inactivity status by imposing a ‘stay home’ and ‘social distancing’ measure as a form of curbing the infection (2, 3). This ‘dual pandemic’ of physical inactivity and COVID-19 has not only affected our activities of daily life (4), but has also resulted in sporting activities of elite athletes to come to a standstill. In particular, the postponement of the Tokyo Olympic/Paralympic Summer games 2020 has severely impacted the global sporting schedules compelling athletes and coaches of sports federations to quickly determine and adopt innovative strategies to augment performance while controlling the spread of COVID-19 infections.

With viral transmission mechanisms becoming clearer, preventive measures have become the mainstay of addressing individual-level risk control (5). However, these actions are sometimes impractical when dealing with elite athletes. High-intensity sports with a mask-on may interfere with normal ventilation and lead to hypoxia (6). Expecting social distancing in a sport that involves physical contact is inherently futile. Therefore, it is imperative to establish a novel, state-of-the-art support protocol that enhances the performance of athletes looking forward to participate in the forthcoming Tokyo Olympic/Paralympic games in 2021. To this end, we studied the modalities of applying cutting-edge technology in the current scenario and the benefits of such approaches in sports medical sciences. As a model sport that could be susceptible to the aforementioned measures for COVID control, we studied handball athletes in this present report.

Handball is an indoor, contact sport in which two teams pass a ball using their hands, aiming to throw at the goal of the opponent team. The sport requires an intermittent repetition of aerobic and anaerobic demands during games, resulting in a high respiratory rate in such athletes (7). The WHO considers such indoor contact ball games as a high-risk sporting activity for virus spread (8). Under the aforementioned playing conditions, if an infected player gets included in the team, it is likely to facilitate the transmission of the virus and to cause vast clusters. Currently for athlete training, by limiting contact with the outside world, the ‘bubble’ style training camps are known to reduce the risk of infections, thereby allowing safe training practices with teammates leading to high compliance (9). Considering the high success rates of ‘bubble’ camps (10) and a fundamental understanding of infection control for COVID-19, we utilized these principles to establish a high-impact safe training/on-court game environment incorporating novel risk-monitoring methodologies to improve athlete performance in contact sports.

The conceptualized control strategies / guidelines in elite sports and sporting events were extensively announced in 2020 (11–13). Specifically, with respect to handball training/playing environments during the COVID era, research is limited. Due to a paucity of literature in this field, we present here a report on the national-level handball team that will participate in the Tokyo-Olympics with these guidelines in consideration. Under the initiative of Osaka University SRIP (Sports Research Innovation Project) supported by JSA (Japan Sports Agency), we assisted the reboot of the Japanese National Women Handball team’s training camp which was unwillingly suspended since December 2019 due to the COVID-19 outbreak. The main purpose therefore was to provide an overview of the challenges encountered during the reboot of high-performance sporting activities of the handball team during the 3rd wave of COVID-19 pandemic in Tokyo, Japan. Our intention of this activity was to understand the requirements of the athlete training camp and provide awareness that coaches, trainers, policy makers from the national sports federations can make use of. Actions that formed the foundation of this reboot were: 1) Web-based health-monitoring, 2) SARS-CoV-2 screening with frequent polymerase chain reaction (PCR) test, 3) Real-time monitoring of social distancing on-court using video-based artificial intelligence (AI) algorithm, and 4) Heart rate and physical intensity evaluation using wearable inertia sensors. This effort was jointly carried out by the Japan Handball Association (JHA), the Osaka University, and the Japan Sports Agency (JSA) leading the activity of JS-CPS (Japan Sports-Cyber Physical System) of Sports Research Innovation Project (SRIP, Japan Sports Agency, Japan).

## 2. Material and Methods

### 2-1. Participant information

We evaluated 29 Japanese national women’s handball players and 24 staff members (including temporary staff) who participated in this camp. The purpose of the study was explained to all participants with documents authorized by the JHA and the ethics committee of the Osaka University Hospital (19537-2). Written informed consent was obtained from all participants. Every athlete was a domestic player at the time of testing. Their home-teams were at Kumamoto, Kagoshima, Ishikawa, Osaka, Mie, Tokyo, Ibaraki and Hiroshima prefectures, and travelled to Tokyo on 24^th^ November 2020 after confirming COVID-19 negative with pre-camp PCR test as described below.

### 2-2. Details of the first domestic training camp after COVID-19 Japan’s state of emergency “Stay-home” social policy

The training camp was held at the Ajinomoto National Training Center (ANTC) in Tokyo. ANTC is a national sports-complex designated for high performance sports. This was the first time a training camp of this scale was organized after COVID-19 Japan’s state of emergency “Stay-home” social policy. The camp started on 24^th^ Nov. 2020 and was completed on 8^th^ Dec. 2020. The reboot of the camp was approved by the Japan Olympic Committee (JOC), the JSA, and the Japanese Ministry of Health, Labor and Welfare (MHLW), based on well-planned regulations within the training halls, as well as emergency protocols prepared by the JHA in advance. On Day-1, all players and staffs were briefed by the JHA with reference to the protocols for team compliance during the training camp.

### 2-3. Scientific monitoring

#### 2-3-1. Web-based health-monitoring

All athletes documented their body temperature, body weight, and subjective health conditions every day via a web-based system (One Tap Sports, Euphoria Co., Ltd., Japan). Monitoring duration was from 10^th^ Nov. to 22^nd^ Dec. 2020 (2 weeks before and after the training camp). When body temperatures above 37.5°C were reported, the system alerted the medical staff via email regarding the athlete’s status.

#### 2-3-2. Screening of SARS-CoV-2 with the frequent PCR test

A total of 10 PCR testing session (364 samples) which included pre-camp screening, in-camp, and post-camp inspection were performed. Saliva samples were examined by SARS-CoV-2 Direct Detection RT-qPCR Kit (Takara Bio, Otsu, Shiga, Japan) using LightCycler 96 (Roche Molecular Systems, Branchburg, NJ) for the detection of SARS-CoV-2 according to the methods described by the manufacturers. Briefly, samples were heated for extraction at 95°C for 5 minutes. For amplification, cycling conditions for heated samples consisted of 52°C for 5 minutes, followed by 95°C for 10 seconds and 45 cycles of 95°C for 5 seconds and 60°C for 30 seconds.

Samples were considered positive when a signal was detected based on the (cycle threshold) Ct value, negative if only the internal control was amplified and invalid when internal control was negative. After completion of the training camp, a self-reported online questionnaire regarding the impression and general impact of PCR testing was carried out among the participants. Their level of agreement was recorded in the form of a 5-point Likert scale.

##### Pre-camp screening and post-camp verification

Each individual sampled their saliva (1mL) at their residence under the online guidance of the researcher (I.O) on 19^th^ Nov. and 9^th^ Dec. thereby providing pre and post-camp samples respectively. The centrifuge-tube (primary packaging) containing saliva was wrapped with an absorbent sheet and was tightly sealed in the Bio-pouch (secondary packaging). The sample was then placed in a bio-mailer box (3rd packaging) and sealed with a security sticker. Finally, the bio-mailer box was put into a 4th packaging box. This box was then transported under the regulation of MHWL (Category B, UN3373) to the Group of Infection Control and Prevention (G-ICP) in Osaka University Hospital via of Japan Post Co., Ltd. service.

##### SARS-CoV-2 PCR testing protocol in the camp at ANTC

During the training period (from 24^th^ Nov. to 8^th^ Dec.), SARS-CoV-2 PCR testing was conducted every Monday, Wednesday, and Friday (ideally every 48h, excluding weekend)(14). On the ‘test day’, each athlete and staff sampled their saliva by about 8:00 am. The samples were then immediately transported to the semi-onsite PCR center held in the Toho University Medical School (about 1.0 hour from ANTC) and tested by the staff of Osaka University ICP team. The results were usually reported to the team until about 14:00 pm, prior to the start of the afternoon training session.

#### 2-3-3. Real-time automated quantitative monitoring of social distancing with motion image-based AI system

The degree of social distancing on the handball court was monitored by a custom-made AI system (Fig. 1). The system calculated the interpersonal distance in real-time using motion images and scored the degree of proximity (Japanese Patent Application No.2020-123769). When the system identified two athletes within a distance of 2.0 m, it assigned a unique proximity ID to that pair, and added one point every second when this distance was maintained continuously. The total sum of the points for all ID pairs normalized by the number of associated athletes was then defined as the cumulative proximity score. The cumulative proximity score was normalized by two separate unit time (UT) scales to consider the effect of respiratory rate (RR) on physical activity, since the risk of virus spread is theoretically proportional to the respiratory flow that is increased due physical activity (15, 16). The UT roughly assumes a time needed for one inhalation-exhalation cycle and was defined, at rest UT = 4 seconds, and at gameplay/training UT = 2 seconds, with the assumption that the ratio between HR and RR is about 4:1 (16). The total time that was maintained continuously for each ID served as a metric for proximity duration.

**Figure-1:**
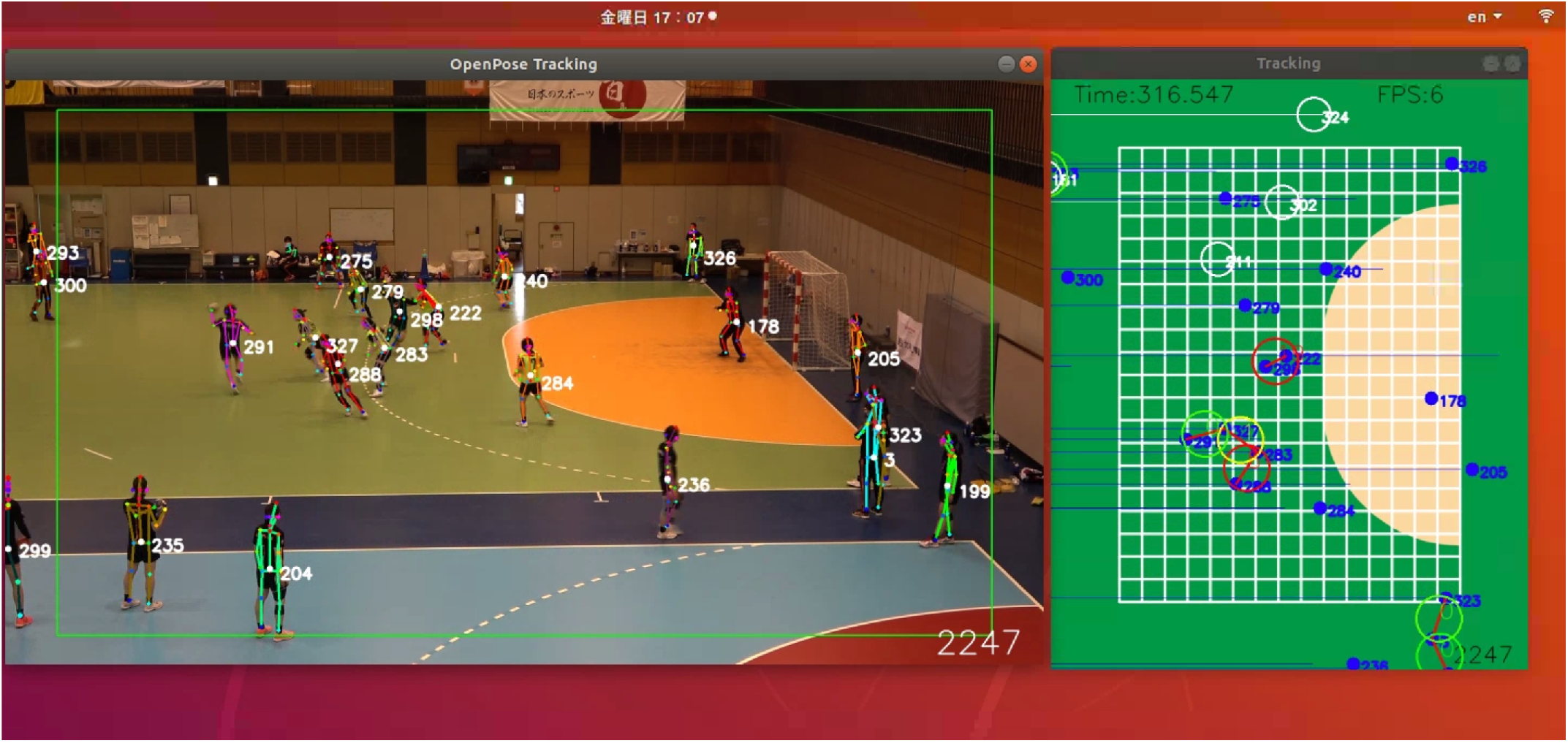
Real-time automated quantitative monitoring of social distancing with motion image-based AI system, shown on Ubuntu 18.04.5 LST. This system identified the participant positions using a function on OpenPose (17). The distance between the players was calibrated based on known court geometry. Once the system identified a pair of athletes within 2m, a white circle appears on the right window. Each additional second, the color of circle turned: green (2 seconds), yellow (3 seconds) and red (>4 seconds).

With the aforementioned metrics in place, we evaluated two training games (each lasting 15 min) held on 6^th^ Dec. 2020. Additionally, midway during the camp (Day-5), the system identified that the stretching time (pre-game) contributed to high proximity scores. We intervened and provided feedback to the teams to disperse during stretching time on the remaining days. Proximity scores are therefore described under results before (up to Day-5) and after (Day-6 onwards) trainers’ feedback.

#### 2-3-4. Heart rate and activity monitoring with wearable sensor device

To quantify the intensity of physical activity, we used an wearable sensor which measured the heart rate (HR) and truncal acceleration. The purpose of this recording was to evaluate whether the physical demands on each player was high enough to enhance their performance. Athletes wore electrodes sewn onto sports-bras with a small wearable sensor (SS-ECG, Teijin Frontier Sensing Co., Ltd, Japan) to record the electrocardiogram (ECG) and three-dimensional acceleration of torso at 200 Hz. 3D accelerometer data was used only to define the active phases during the training matches. The measurements were performed during a 3,000m run (a high intensity endurance test, 30^th^ Nov) and on two training matches (15 min, 6^th^ Dec.).

#### Statistical analysis

The scope of this study was to report an observation of the training patterns and challenges encountered during the pandemic. As a result, no formal sample size calculations were performed. Statistical evaluation included descriptive measurements of key variables of interest; raw data analysis and graphing were performed either on Matlab R2020a and R software v4.0.2. Data analyzed in this manuscript will be made available from the corresponding author upon reasonable request.

## 3. Results

Demographic information of the participants is detailed in Table 1.

**Table 1:** Demographic information of the participants.

|  | **Athletes** | **Staff** |
| --- | --- | --- |
| Number (male) | 29 (0) | 24 (20) |
| Mean age (SD) | 28.4 (4.1) y | 40.2 (14.2) y |
| Mean height (SD) | 163.8 (18.8) cm | 171.2 (11.1) cm |
| Mean weight (SD) | 66.7 (8.3) kg | 70.7 (13.2) kg |
| History of COVID-19 infection before camp | None | None |
| COVID-19 infections after camp | None | None |

The reporting rate of the web-based monitoring was 88.7 % throughout the survey period (10^th^ Nov. to 22^nd^ Dec.) and 96.6 % during camp (24^th^ Nov. to 8^th^ Dec.). Every athlete completed their web-based monitoring.

SARS-CoV-2 PCR testing history and results are summarized in Table 2. According to the results of pre-camp screening (all negative for SARS-CoV-2, informed via email), athletes were invited to the training camp by the JHA. In-camp PCR among all athletes was also negative. The mean sample-to-answer (turnaround) time was 4.2 hours (including 1.0 hour of sample transportation time), therefore the team staff could announce the result of PCR test around 13:00 on every testing day, prior to the start of the afternoon training session (∼15:00). To shorten the turnaround time, we launched a dedicated PCR center near ANTC. Following the training camp, results of the post-camp screening (all negative, also informed via email) were reported to JHA on 10^th^ Dec.

**Table 2:** PCR test history and results.

| Date | Schedule | Samples: Athletes | Samples: Staff | Results | Sample-to-answer time* | Note |
| --- | --- | --- | --- | --- | --- | --- |
| 20<sup>th</sup> Nov. | Pre-camp | 28 | 21 | Negative | - | All athletes and staff transported their saliva samples to Osaka University from their home team. Based on this result, JHA recruited all athletes and staffs. |
| 24<sup>th</sup> Nov. | In-camp, start up | 28 | 11 | Negative | 5.5 h |  |
| 25<sup>th</sup> Nov. | In-camp, regular | 28 | 10 | Negative | 3 h |  |
| 25<sup>th</sup> Nov. | In-camp, irregular | 1 | 0 | Negative | 3 h | One athlete was additionally invited. |
| 26<sup>th</sup> Nov. | In-camp, irregular | 4 | 0 | Negative | 3 h | One athlete got fever on 26<sup>th</sup> morning. This athlete as well as three athletes from the same home team were tested to screen. |
| 27<sup>th</sup> Nov. | In-camp, regular | 29 | 12 | Negative | 5.5 h |  |
| 30<sup>th</sup> Nov. | In-camp, regular | 29 | 12 | Negative | 5 h |  |
| 2<sup>nd</sup> Dec. | In-camp, regular | 28 | 11 | Negative | 5 h | One athlete retired due to injury. |
| 4<sup>th</sup> Dec. | In-camp, regular | 28 | 10 | Negative | 4 h |  |
| 7<sup>th</sup> Dec. | In-camp, regular | 28 | 11 | Negative | 4 h |  |
| 10<sup>th</sup> Dec. | Post-camp | 29 | 6 | Negative | - | All athletes and staffs transported their saliva samples to Osaka University from their home team. Based on this result, JHA allowed them to return to the regular practice and job. |

\* Sample-to-answer time or turnaround time was the time from the sampling of athletes' saliva to the arrival of result e-mail. This time included both the sample transportation time (approximately 1.0 h) and the inspection time. Our PCR test center (Toho University Medical School) was 28 km apart from the training hall (ANTC) thus approximately 1.0 h by a car.

A summary overview of the temperature checks documented by the participants is graphed in Fig. 2. As shown, a single athlete reported febrile status on Day-3 (∼38.5°C). According to the emergency protocol made by the JHA, all athletes were temporarily quarantined until the PCR results were confirmed. An exclusive PCR test for this febrile athlete was performed. Three other athletes from the same home-team were also tested immediately. Though this was not the regular ‘test day’, samples from these four athletes were dispatched at 10:00 and the results (all negative) arrived by 13:00. On confirmation of a negative result, all athletes except the febrile athlete restarted their training schedule on the same day. The sick athlete returned to the training after recuperation on 3^rd^ Dec. with the 3 additional PCR negatives.

**Figure-2:**
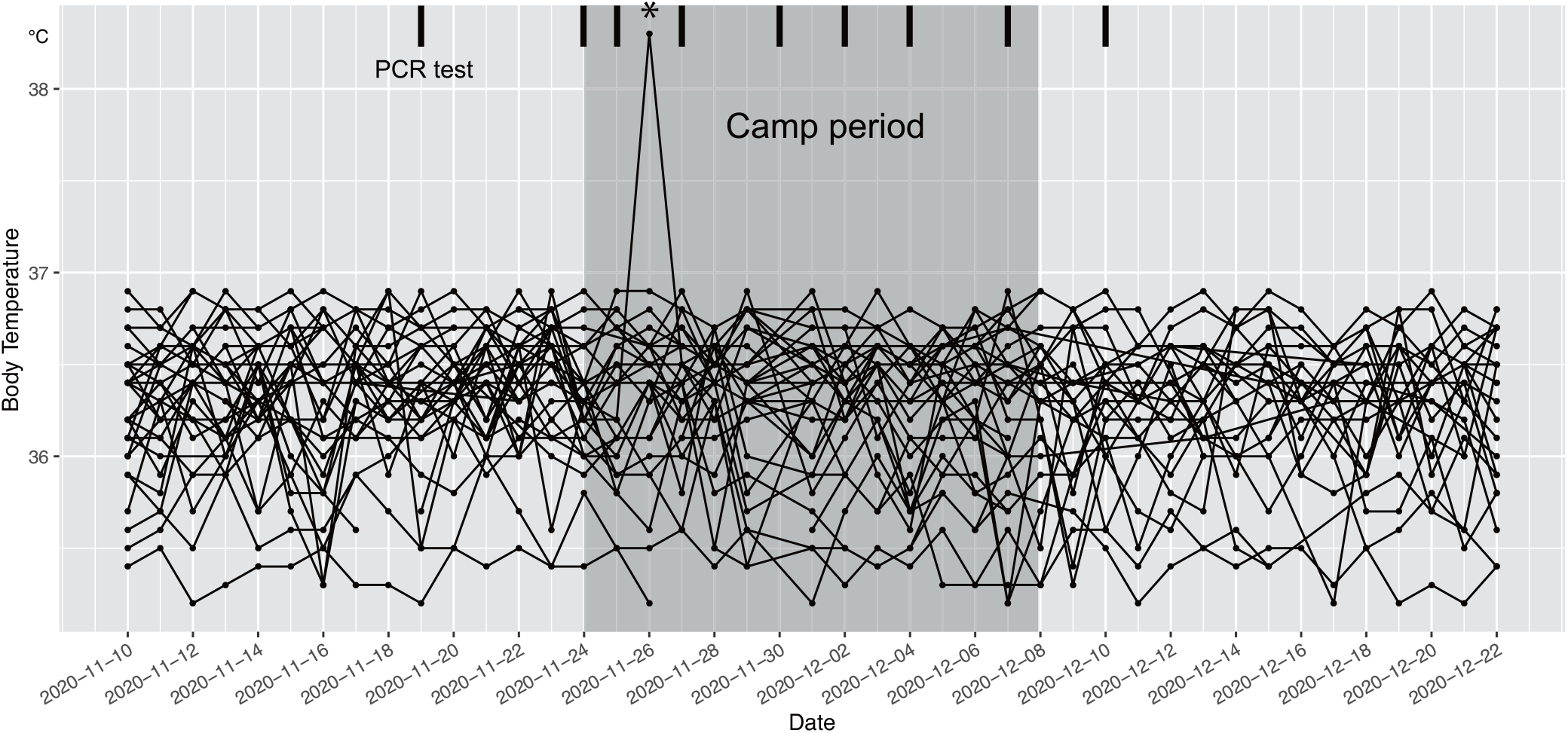
Body temperature chart line-scatter plot for all athletes. Grey shaded area is the camp period (15 days), thick vertical lines denotes the day PCR tests performed, Asterisk denotes the single febrile athlete recorded on Day-3

Results of the online questionnaire about infection test protocol were obtained from 27 athletes and 10 staffs (Fig. 3 Q1-7). Our survey suggested that most participants felt that 3 tests a week was too frequent (Q1) and were not compliant (Q2), but recognized the importance of high frequency testing to maintain a safe training environment (Q3). The frequent testing with a short sample-to-answer time provided a strong sense of relief both to athletes and staff to participate in the handball training sessions (Q4, 5). Moreover, being aware of the negative results following every testing session increased their compliance to report minor symptoms (such as coughing) to the medical staff (Q6). The most of members felt anxious while waiting for the result each test day (9 strongly agree and 16 agree), however, most appeared unconcerned about the PCR results (5 neutrals and 7 disagree, Q7).

**Figure-3.**
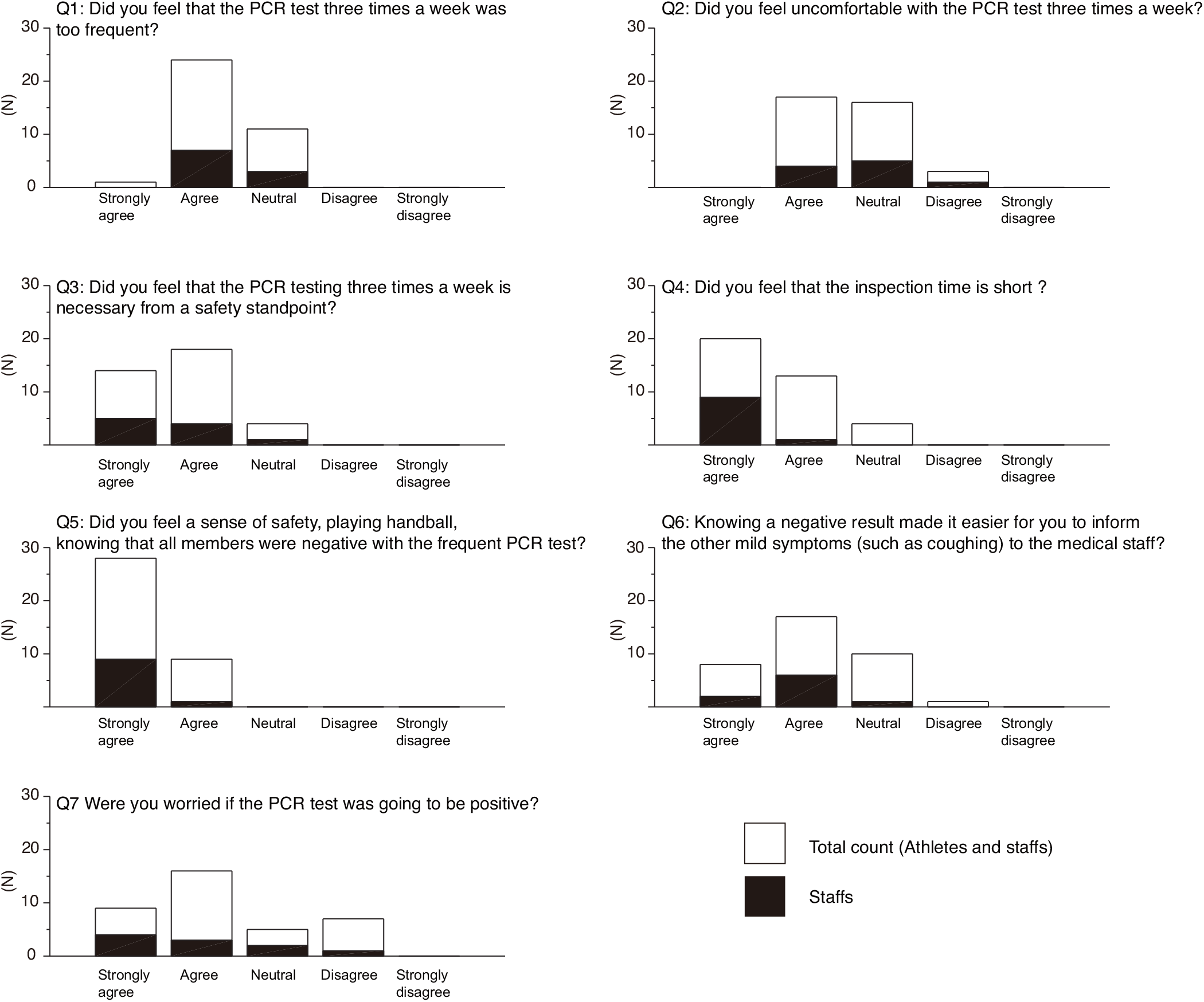
Results of online questionnaire regarding perception of PCR tests during the camp.

Table 3 describes the situation specific proximity scores gathered from real-time video-based AI monitoring. During the 15-min training matches, the cumulative proximity scores were 10.4 and 17.2. According to the Japanese guidelines (18), the definition of close contact is the distance within 1 m maintained for 15-min. The cumulative proximity score of this definition is 15 min * 60 seconds / 2 seconds of UT / 2 persons = 225 points. Therefore, it was found that the cumulative proximity score during 15-min training matches were lower than the Japanese definition of the close contact. Although a high proximity occurred along the goal-area line (Fig. 4), the histogram of continuous time for each proximity ID illustrated that during the match, the frequency of short-duration IDs was high, while during stretching, the frequency of long-duration IDs was high, because every player was dynamically moving during the games (Fig. 5).

**Table 3:**
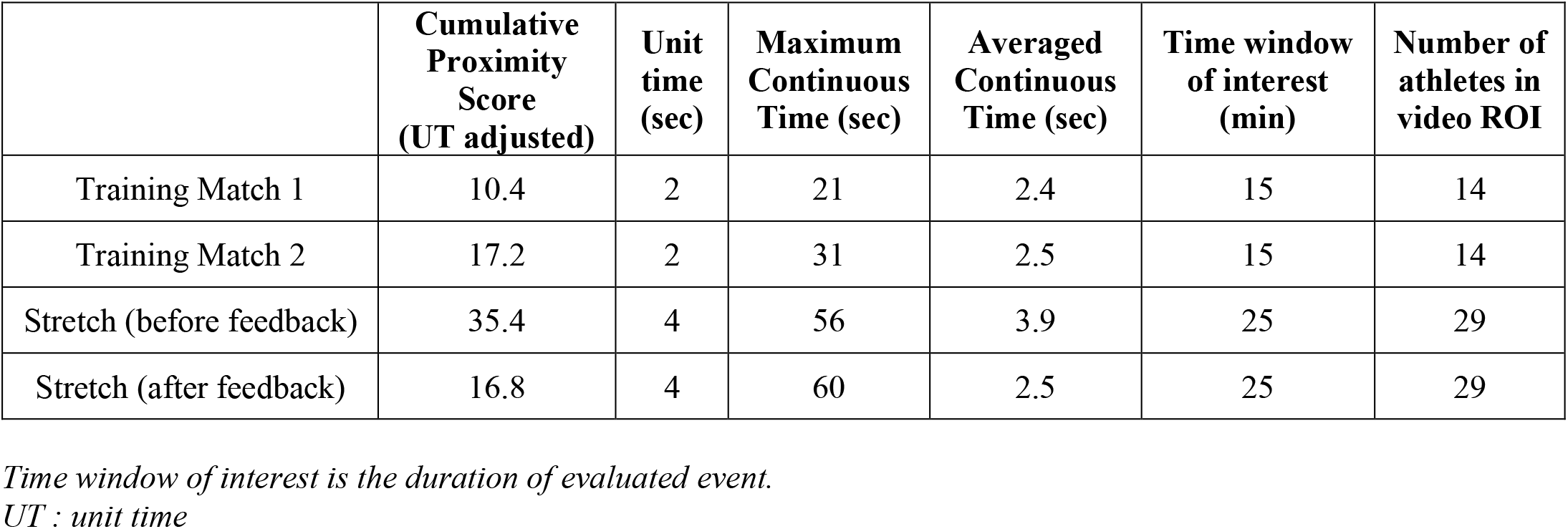
Situation specific proximity scores.

**Figure-4:**
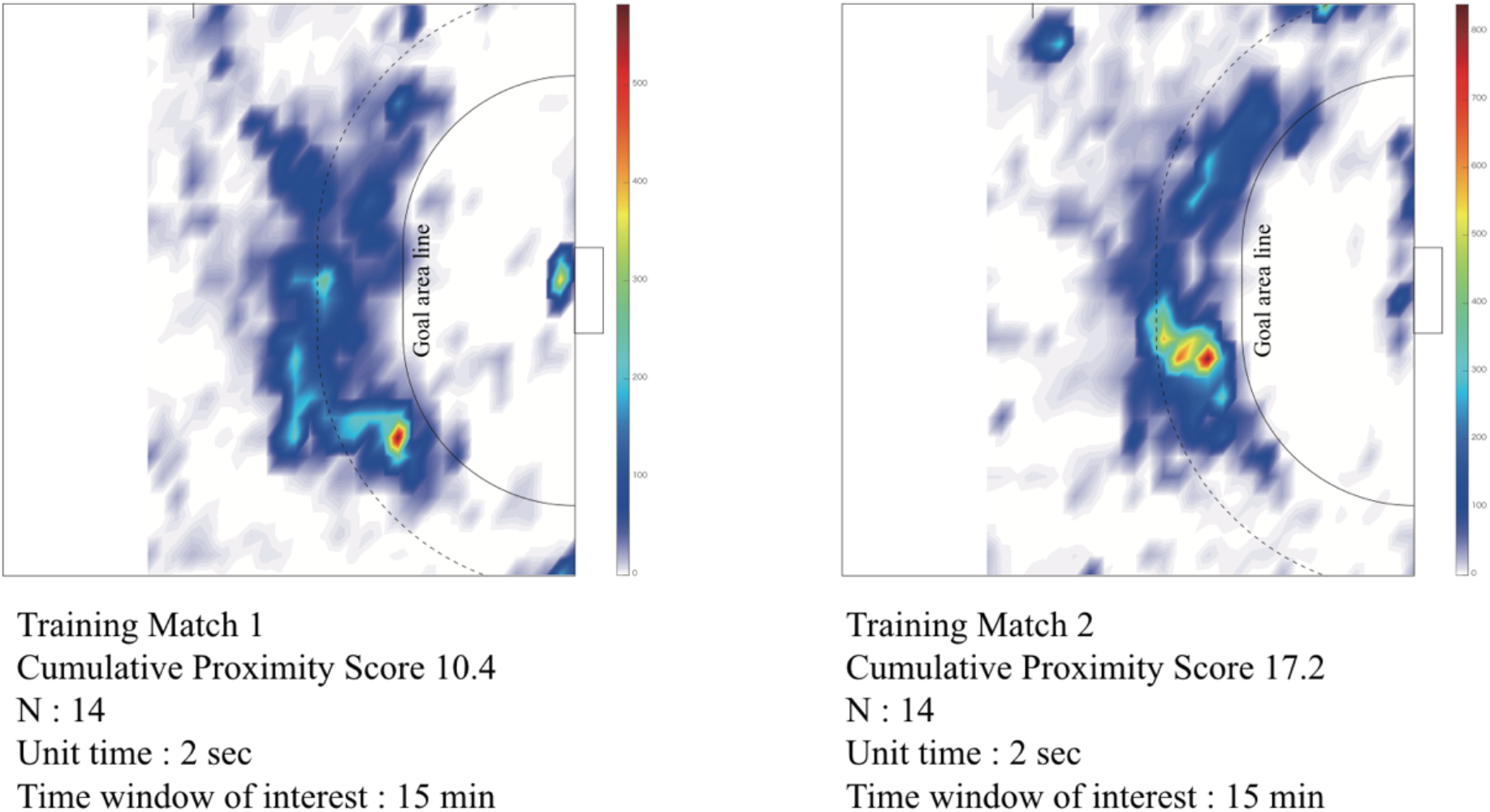
Heat map of area specific proximity density on the handball court during training match

**Figure-5:**
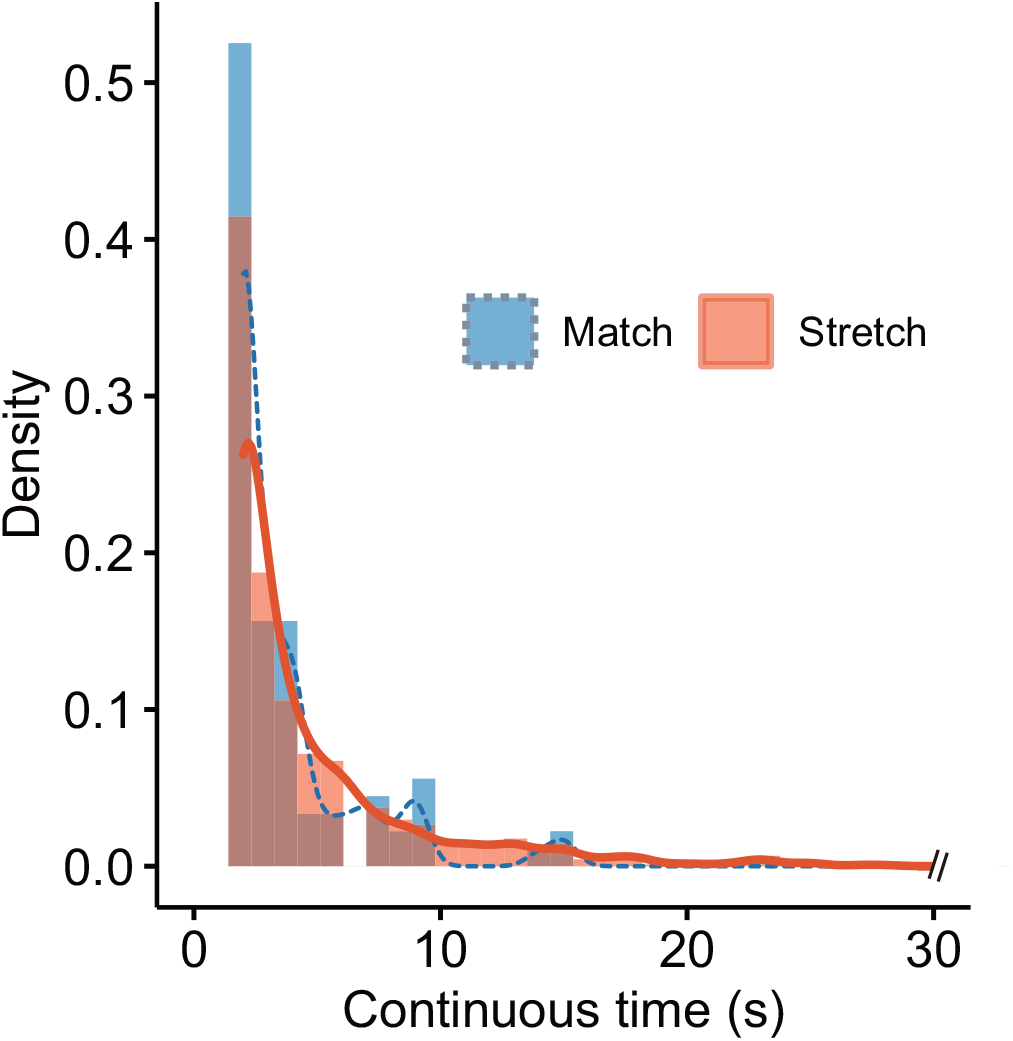
Density plot of the duration for each proximity ID during training and stretching time.

We found that many athletes concentrated on a specific place during stretching time before practice (score = 35.4, averaged continuous time = 3.9 seconds). After providing feedback on this issue to the team on 27^th^ Nov. (Day-5), the location of the athletes during stretching were dispersed and proximity condition was clearly reduced thereafter (score=16.8, averaged continuous time = 2.5 seconds, Fig. 6).

**Figure-6:**
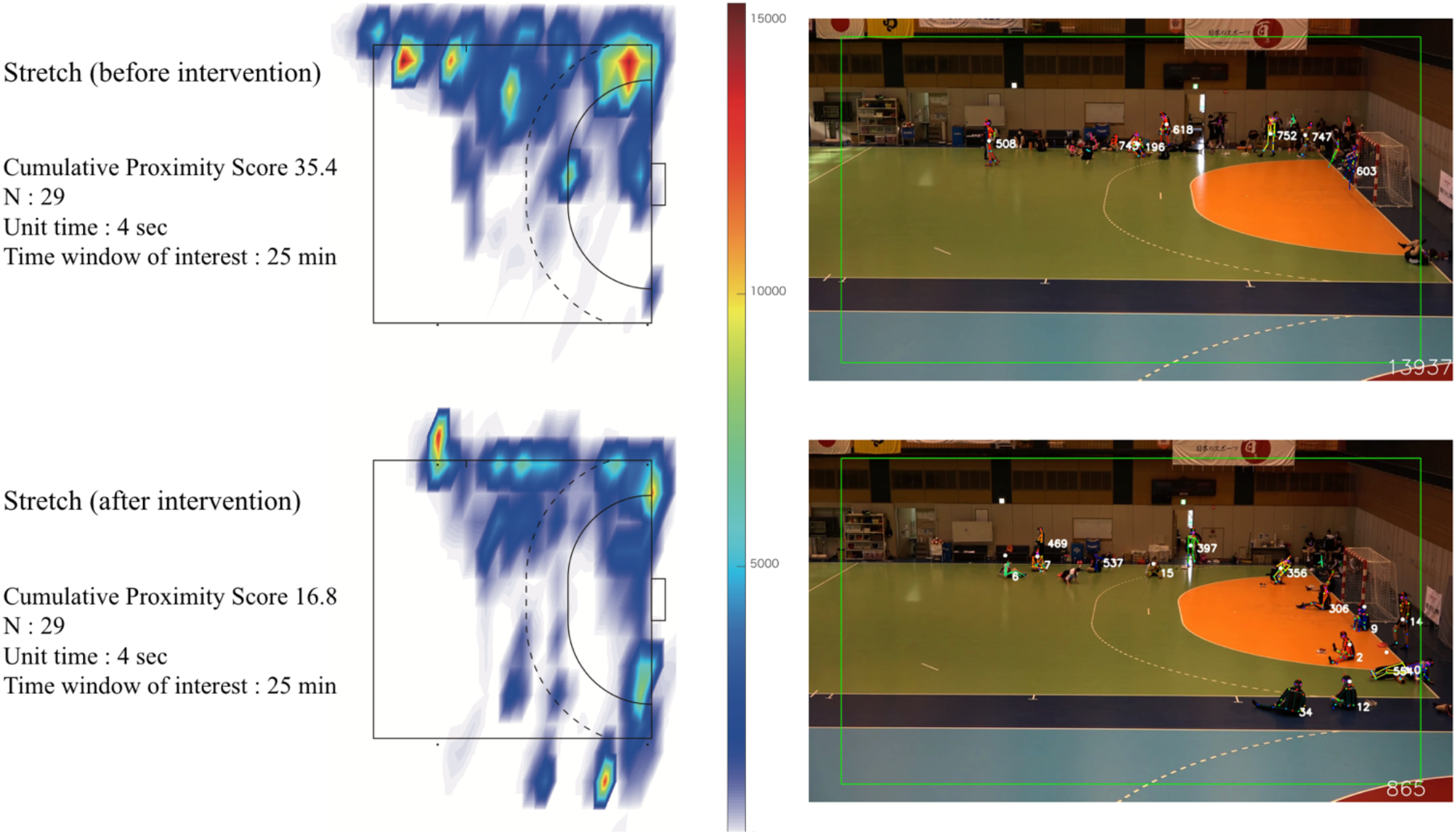
Transition of high proximity distribution during stretching, before and after intervention.

Evaluation results of HR and physical endurance is shown in (Table 4). During the training matches, the averaged maximum HR achieved by pivot player (PV) = 192.2 bpm and by back player (BP) = 188.1 bpm. These players have the most active roles on the court. As reference to the HR value measured during the 3,000m endurance run (a vigorous exhausting running test), the maximum in-match %HR by BP and PV scored more than 100% of 3,000 m run. However, the averaged in-match %HR ranged between 67.6% (Goalkeeper) and 78.5% (BP).

**Table 4:**
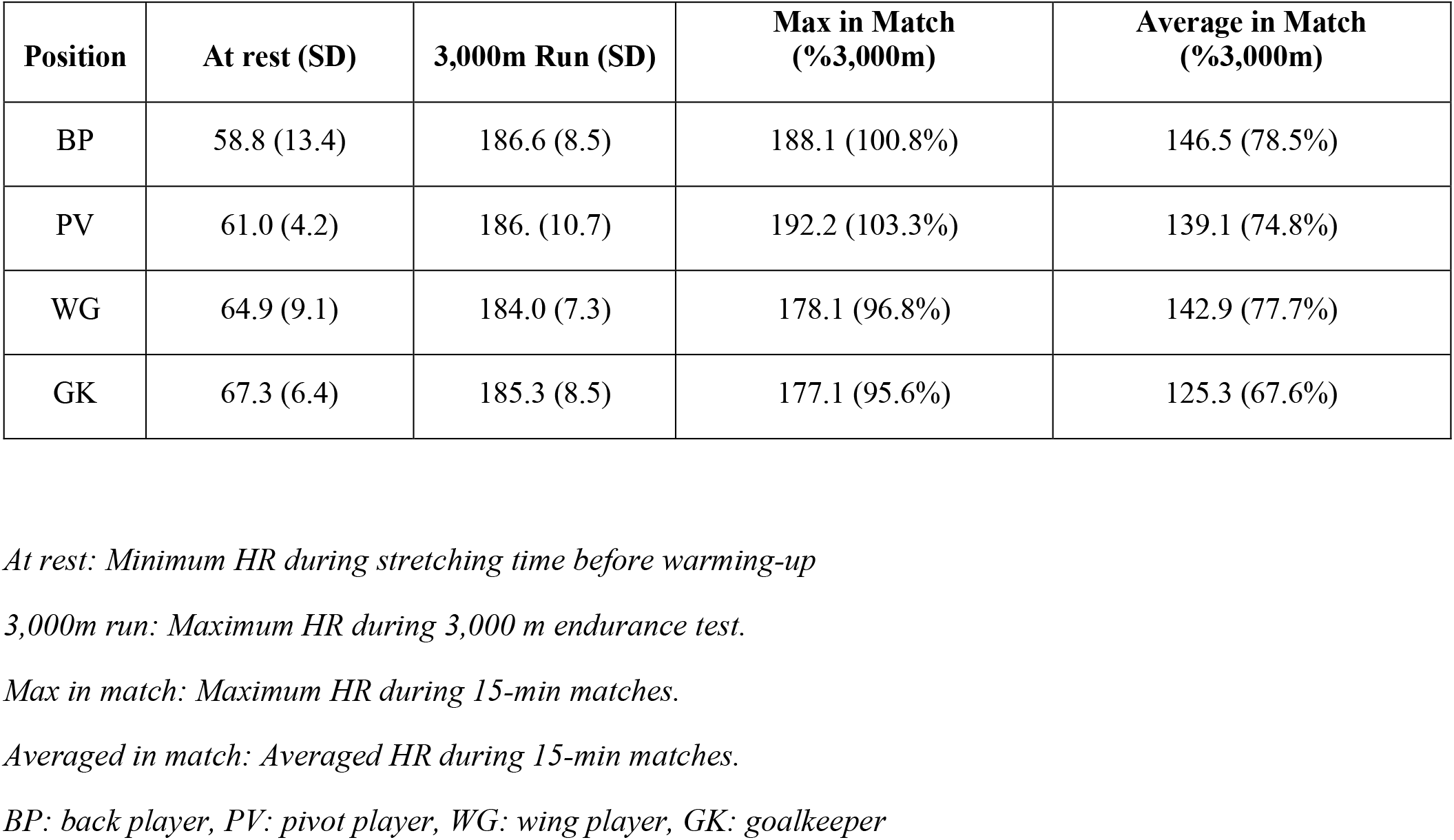
Heart rates at rest, during 3,000m endurance run, and training matches.

| Position | At rest (SD) | 3,000m Run (SD) | Max in Match (%3,000m) | Average in Match (%3,000m) |
| --- | --- | --- | --- | --- |
| BP | 58.8 (13.4) | 186.6 (8.5) | 188.1 (100.8%) | 146.5 (78.5%) |
| PV | 61.0 (4.2) | 186. (10.7) | 192.2 (103.3%) | 139.1 (74.8%) |
| WG | 64.9 (9.1) | 184.0 (7.3) | 178.1 (96.8%) | 142.9 (77.7%) |
| GK | 67.3 (6.4) | 185.3 (8.5) | 177.1 (95.6%) | 125.3 (67.6%) |

*At rest: Minimum HR during stretching time before warming-up*
*3,000m run: Maximum HR during 3,000 m endurance test.*
*Max in match: Maximum HR during 15-min matches.*
*Averaged in match: Averaged HR during 15-min matches.*
*BP: back player, PV: pivot player, WG: wing player, GK: goalkeeper*

## 4. Discussion

We present here an overview of the challenges encountered by the JHA in developing a safe training environment and evaluate the outcome of risk management among high-performance athletes who rebooted their training despite the COVID-19 pandemic. We were successful in completing the training camp with no COVID-19 positive players as well as achieving an acceptable physical intensity required to enhance their handball performance with this comprehensive monitoring/feedback protocol.

### 4-1. Web-based health-monitoring

Continuous monitoring of an accurate health status is one of the first steps to minimize the spread of infection in a closed community such as a training camp. We observed no severe health problems related to the current training protocol despite one athlete presenting with fever (38.5 °C) on the third day of the training camp (Fig. 2). With the help of web-based system, the medical staff could immediately observe and identify health concerns when the system reported any issues. This also enabled us to manage all players’ health status while avoiding unnecessary contact between them.

A possible limitation of this system was that the responsibility of data entry rested solely on the athletes. There was a fear of missing out on competitive opportunities making athletes occasionally hesitant to disclose their health status. While it is commendable of the athlete who reported her fever, it was important to build a sense of safety among other athletes as well. We surmised that crucial appraisal regarding infection control by the JHA and frequent negative PCR testing promoted a positive mental outlook to genuinely report their true health condition (Fig. 3, Q: 5, 6). Regardless, web-based monitoring contributed positively towards developing a conducive behavioral environment among the athletes.

### 4-2. SARS-CoV-2 PCR testing and feedback

Though we had no SARS-CoV-2 positive athletes during the entire duration of the camp, our frequent testing schedule (every 48h or 72h) enabled us to detect COVID-19 infectious athlete early, thereby minimizing the risk of virus-spread. Larremore et. al. suggested that repeated screening of susceptible but asymptomatic individuals could be used to limit transmission and its effectivity therefore rests on accessibility, frequency and sample-to-answer time (Larremore *et al*., 2020). The accessibility of the PCR test turned out to be crucial when an athlete was reportedly febrile. Though it was outside of the ‘testing day’, the prompt PCR testing allowed us to confirm a ‘negative’ test within 3-hours of sample collection. Without the convenience of obtaining a PCR result in the backdrop of a febrile patient, this would have necessitated a complete shut-down and quarantine of the training camp irrespective of the actual infection status. Due to a well-planned emergency protocol in force, we could minimize the training loss, allowing the team to return to their regular training schedule on (the afternoon of) the same day.

The results of the online PCR survey highlighted the increased compliance of the athletes despite the high frequency of testing (Fig. 3). While the participants were aware of the risk of transmission in a closed set-up like this, the additional burden of testing did not, in fact, hamper their training sessions or their main objective in the camp i.e. enhancing performance. Considering the effectivity of PCR testing in such scenarios, we recommend that for national training centers such as ANTC, an exclusive PCR center as a primary facility will be indispensable in the future. Furthermore, it would be desirable to establish a mass screening methodology that enables a low-cost on-court inspection to improve test accessibility.

### 4-3. Real-time automated qualitative monitoring of social distancing on the handball court using video-based AI algorithm

Real-time quantitative monitoring using AI demonstrated the characteristics of the proximity status that occurred during on-court handball training. We observed that the daily routines for all sports teams, such as stretching exercise on the floor before and after practice, cause significant proximity. Even minor feedbacks resulted in small behavioral changes (such as increasing interpersonal distance during stretching), substantially reducing the proximity scores. In this way, the AI system was able to provide objective numerical scores to improve athlete behavior.

If the stretching time could be construed as ‘control’ environments, the in-game matches served as ‘test’ conditions to compare the proximity status between the players. The continuous-time density plot revealed that the proximities during a handball match were not as long-lasting as it was during stretching (Fig. 5). The cumulative proximity scores during handball matches were considerably lower when compared to those during stretching (Table 3). Due to the highly dynamic nature of the sport, it was difficult to conclude the proximity outcomes in matches since none of the players were near-enough and/or long-enough for significant change in the readings. Conversely, we could interpret that despite being a body-contact sport, the athletes are never too close for long durations which is always a risk for virus-transmission. Despite being a contact sport, there appears to be low evidence of transmission to limit sporting activities (19). Without excessive speculation, we suggest that if the camp is ‘bubbled’ with players who have tested negative, the results of real-time AI monitoring systems can positively influence the decision to remove masks, allowing athletes to play handball normally.

### 4-4. Heart rate and physical intensity evaluation using the wearable sensor

Since the primary aim of training camp was to increase athlete performance, it was essential to quantify whether the physical intensity was demanding enough to fulfill this objective. Kniubaite et al. reported that the averaged %HR of the elite Lithuanian female handball players during international competition was from 84.2 to 84.8%. Póvoas et al. also evaluated the in-match %HR of the male players (Portuguese handball professional league) and reported that the averaged %HR was 83%. In comparison, the averaged %HR of 74.8—78.5% in Japanese players (BP, PV and WG) were slightly lower than that of prior reports (20, 21). Despite the differences in analytical methodologies (official competitions vs training matches), the physical demands on the Japanese athletes experienced in the training matches were nearly comparable to those during competitions. Within reason, we speculate that the aforementioned training in the camp was high enough to allow substantial enhancement in the athletes’ performance despite the stringent infection control protocols.

### 4-5. Limitations

Whereas the rapid and flexible PCR testing procedure minimized the impact on the players’ training schedule, it is important to note that the sample size for our study was low (29 athletes and about 10 staffs per test day). Although our presented methodologies could be optimum for small sample sizes, for large-scale screening (>1000 samples) in the future, the person organizing the team should consider an optimal screening strategy in terms of cost, technician availability, testing facilities, and warranting the test accuracy. The video-based position detection is a vital technology for large indoor environments where GPS or other positioning systems do not work. Since the technology is new, more comparative data is needed to fully validate its efficacy on the risk quantification measured via proximity scoring. In its current state, we have provided a tentative interpretation of what high/low cumulative proximity scores are to be expected for infection control. In addition to increasing sample sizes, it will be beneficial in the future to implement individual assessment features to the current cumulative evaluation.

## 5. Conclusion and future directions

Aiding athletes in disrupting physical inactivity and normalizing their training routines during the pandemic was our central purpose that led to the rebooting of high-performance sporting activities under JS-CPS (Japan Sports-Cyber Physical System) and by the SRIP (Sports Research Innovation Project). The pursuit resulted in the successful management of a training schedule among handball players culminating in decent improvements in their performance within a stringent COVID-19 screening environment. The sum of our efforts in this current work is expected to set the foundation for future endeavors in standardizing training protocols and, in turn, scaling up to include for other team-based indoor sporting activities. Specifically, we believe that the short turnaround time for PCR testing is vital for the early identification and prioritization during the screening process. As an added benefit, the outcomes of this process also develop a sense of security among the athletes. Furthermore, an on-site low-cost mass screening methodology will be beneficial to develop in the future. Using information technology, real-time quantification of health status and proximity metrics will be an indispensable tool to quantify the effect of risk management. These technological innovations developed under the SRIP research can be a promising approach in building a safe environment to support athletic training during the pandemic.

## Data Availability

Data analyzed in this manuscript will be made available from the corresponding author upon reasonable request.

## Acronyms

SARS-CoV-2: severe acute respiratory syndrome coronavirus 2
CPS: Cyber physical system
PCR: Polymerase chain reaction test
AI: Artificial intelligence
JHA: Japan Handball Association
SRIP: Sports Research Innovation Project, Osaka University
ANTC: Ajinomoto National Training Center
JSA: Japan Sports Agency
JOC: Japan Olympic Committee
MHWL: Japanese Ministry of Health, Labor and Welfare
G-ICP: Group of Infection Control and Prevention

## Data availability statement

Data that support the findings of this study will be made available from the corresponding author upon reasonable request.

## Funding

This work was supported by the Sports Research Innovation Project (SRIP) grant, sponsored by the Japan Sports Agency.

## Acknowledgement

We appreciate a great cooperation of the Japanese women’s national handball team (Orihime Japan) players and associated staffs Ulrik Kirkely, Antoni Parecki, Ryosuke Kushida, Yosuke Kakazu, Minako Iwatani, Ai Fujita, Kosuke Kaya, Yuta Yoshimura, Manabu Todoroki, and Chiaki Kawakami.

## Contributions

I.O., S.H., Y.A., N.O., K.T., and K.N. conceptualized and designed the study.

I.O., T.T., K.I., N.O., and K.S. organized the experiments on during the camp.

I.O., S.K., and R.H. performed the experiments and data collection.

S.H., Y.A., N.O., and K.T. performed PCR test.

R.H., A.U., and T.H. developed artificial intelligence proximity detection system.

I.O., S.K., A.U., and R.H. analyzed the data and performed the statistical analysis.

I.O., G.S.R., and K.N. wrote the manuscript.

I.O., G.S.R., K.Y., T.H., and K.N. reviewed the manuscript, suggested corrections and approved its final version.

## Notes

### Competing Interest Statement

The authors have declared no competing interest.

### Author Declarations

This study was authorized by the Japan Handball Association and the ethics committee of the Osaka University Hospital (19537-2)

